# Metabolism of the antioxidant micronutrient ergothioneine as a plasma biomarker of cognitive resilience in older people with Alzheimer’s disease amyloid pathology

**DOI:** 10.1101/2025.07.31.25332054

**Authors:** Joyce R. Chong, Irwin K. Cheah, Richard M. Tang, Barry Halliwell, Christopher P. Chen, Mitchell K.P. Lai

**Author notes:** **Corresponding Author:** Mitchell K.P. Lai, PhD, Department of Pharmacology, Yong Loo Lin School of Medicine, National University of Singapore, Unit 09-01, Centre for Translational Medicine (MD6), 14 Medical Drive, Singapore, 117599 Singapore.

## Abstract

**Background:** Associations between dietary micronutrients and cognitive resilience to Alzheimer’s disease (AD) amyloid pathology is currently unknown. We investigated whether plasma levels of L-ergothioneine (ET), its metabolite L-hercynine (HC), and their ratio (HC:ET, as an index of ET metabolism) affect known associations between biomarkers of amyloid pathology (p-Tau181 or p-Tau217) and cognitive decline.

**Methods:** 259 initially dementia-free participants recruited from memory clinics and the community in Singapore had baseline measurements of plasma p-Tau, ET, HC, as well as annual neuropsychological assessments for up to 5 years to derive cognitive trajectories based on Clinical Dementia Rating-Sum of Boxes (CDR-SB) slopes.

**Results:** High HC:ET attenuated the positive correlations between plasma p-Tau and CDR-SB slopes. Compared with participants with low amyloid burden, participants with high amyloid burden had higher risk of cognitive decline when HC:ET was low (Hazard ratio [HR] = 2.33, p = 0.002), but not when HC:ET was high (HR = 1.47, p = 0.32).

**Conclusion:** The identification of ET metabolism as a novel biomarker of cognitive resilience supports further investigations into mechanisms underlying its neuroprotectant properties. ET should be further assessed as a potential candidate for countering amyloid pathology-associated cognitive decline.

## 1. Introduction

Although the hallmark pathological features of Alzheimer’s disease (AD) amyloid plaques and neurofibrillary tangles are closely associated with cognitive decline and dementia, some individuals remain cognitively intact despite substantial pathological burden [1]. This observed discrepancy between severity of brain pathology and cognitive trajectory gave rise to the concept of cognitive resilience, which refers to the capacity of the brain to maintain cognition and function with aging and disease [2, 3]. In the context of AD, cognitive resilience may be operationalized as the multiple adapted or innate processes at the molecular, cellular and network levels which facilitate better-than-expected cognitive function despite the severity of concomitant amyloid pathology in some individuals. While initial studies have investigated sociobehavioral, neuroimaging and genetic factors of cognitive resilience [4], the putative biological mechanisms underlying cognitive resilience have more recently begun receiving research attention, with reports suggesting involvement of neuroinflammatory regulation, neurogenesis, cellular repair, mitochondrial, vascular and synaptic functions [5–7]. In contrast, there is a dearth of studies directly investigating oxidative stress-associated markers in cognitive resilience. Considering the established pathogenic roles played by oxidative stress in both the neuropathologic hallmarks of amyloid plaques and neurofibrillary tangles, as well as in the frequently concomitant features of chronic neuroinflammation and cerebrovascular disease (CeVD) [8–12], it is worth investigating whether the regulation and balance between pro-oxidant and antioxidant mediators or processes may underlie cognitive resilience versus decline in AD. L-Ergothioneine (ET), a diet-derived thione found in high abundance in mushrooms and other foods, has originally been characterized as a potent antioxidant [13] whose effects are mediated via oxidization by reactive oxygen species (ROS), thus rendering ET an important scavenger of ROS [14, 15]. Studies have shown L-hercynine (HC) to be a metabolite of ET in humans [16] produced from the desulfurization of ET by ROS, potentially via the formation of sulfonated intermediates [15, 17]. We have previously reported that deficits in plasma levels of ET correlated with both neurodegeneration and CeVD in participants with dementia [18]. However, whether changes of ET and associated biomarkers may mediate cognitive resilience to amyloid pathology are at present unknown. In this biomarker study, we assessed plasma ET, its metabolite L-hercynine (HC), and their ratio (HC:ET) in a cohort of longitudinally assessed participants who were dementia-free at baseline, and investigated their effects on the established relationship between well-established plasma biomarkers of brain amyloid burden and cognitive decline.

## 2. Methods

### 2.1. Study cohort and clinical assessments

Detailed descriptions of the study cohort have been previously reported [19]. Briefly, the 259 dementia-free participants for this study were recruited from memory clinics and the community in Singapore from August 2010 to July 2019 as part of a larger longitudinal cohort study of dementia (n = 700, see **Supplementary Figure 1**). Determination of clinical risk factors and covariates was based on a detailed questionnaire administered to all participants to collect demographic information and medical history. Education level was defined as years of formal education. Risk factors for concomitant cerebrovascular diseases, such as hypertension, hyperlipidaemia, diabetes mellitus, and cardiovascular diseases were ascertained from clinical interviews and medical record searches, and classified as present or absent. Hypertension was defined as systolic blood pressure of 140 mm Hg or more and/or diastolic blood pressure 90 mmHg or more, or history of antihypertensive medication use. Diabetes mellitus was defined as glycated haemoglobin (HbA1c) of 6.5% or more, or on diabetic medication. Hyperlipidemia was defined as total cholesterol levels of 4.14 mM or more, or on lipid lowering medication. Cardiovascular disease was classified as a previous history of atrial fibrillation, congestive heart failure, and/or myocardial infarction. Carrier status of the Apolipoprotein E gene variant *APOE4*, an established AD risk factor, was defined as having at least one ε4 allele using a standardized genotyping protocol [20]. Participants were also administered a comprehensive battery of neuropsychological tests which assesses seven cognitive domains (see **Supplementary Table 1**) for categorization into clinical subgroups: No Cognitive Impairment (NCI) if participants did not demonstrate objective cognitive impairment on the battery at baseline; while Cognitive Impairment No Dementia (CIND) was determined by the presence of impairment in one or more cognitive domains (defined by a score of at least 1.5 standard deviations below established education-adjusted cut-off values on any component test) at baseline, without loss of independent daily function, and not fulfilling the criteria of dementia diagnosis based on the Diagnostic and Statistical Manual of Mental Disorders, 4th Edition (DSM-IV). Additionally, Clinical Dementia Rating (CDR) assessments [21] as well as a neuropsychological battery which assesses six cognitive domains (executive function, attention, language, visuospatial function, visuomotor speed, and memory) were performed at baseline and annual follow-up visits for up to 5 years.

### 2.2. Cognitive trajectories and secondary outcomes

For cognitive trajectories, the primary outcome measure was CDR Sum of Boxes (CDR-SB) [22] slopes, defined as the annual change in CDR-SB. Of the 700 participants recruited between August 2010 and August 2020, 649 had Clinical Dementia Rating Sum of Boxes (CDR-SB) recorded at baseline as well as at least one annual follow-up visit (mean ± SD follow-up duration of 49 ± 16 months). These participants were included in the calculation of CDR-SB slopes. To extract the CDR-SB slope for each participant, linear mixed effects model modelling CDR-SB over time in the overall cohort (NCI [n=132] + CIND [n=271] + Dementia [n=246]; n=649), with random intercepts and slopes, was used. The median [IQR] annual change in CDR-SB was 0.045 [0.4] for dementia-free participants (n=403) and 1.24 [1.2] for dementia participants (n=246). The secondary outcomes were: (a) Risk of incident cognitive decline, defined by ≥0.5 increment in CDR Global Score (CDR-GS) [23] at any annual follow-up compared to baseline; and (b) Longitudinal trajectories of cognitive decline measured by global cognition and domain-specific cognitive tests on the neuropsychological battery. As this study focuses on cognitive resilience, only dementia-free (NCI and CIND) participants at baseline were included (see participant flowchart in **Supplementary Figure 1**).

### 2.3. Plasma p-Tau measurements and references ranges for amyloid pathology

Details on blood collection, processing and storage were previously published [24]. In this study, we used plasma p-Tau181 and p-Tau217 as established markers of brain amyloid pathology in both western and Asian cohorts [24–27]. Plasma p-Tau181 was performed on the Quanterix Simoa® HD-1 analyzer using commercially available P-Tau181 Advantage V2 assay kits (Quanterix Corporation, Cambridge, MA, USA), per manufacturer’s protocol. Plasma p-tau 217 was measured on a NUcleic acid Linked Immuno-Sandwich Assay (NULISA) platform [28] using the commercially available NULISASeq™ CNS Disease Panel (Alamar Biosciences, Fremont, CA, USA). Raw data were normalized using internal and inter-plate controls, rescaled, and log-2 transformed to derive NULISA Protein Quantification (NPQ) units. Reference ranges for brain amyloid positron emission tomography (PET) positivity was determined by a three-range cut-off approach based on participants within the local cohort with plasma p-Tau181, p-Tau217 and amyloid PET data available (see **Supplementary Figure 2**).

### 2.4. Plasma L-ergothioneine (ET) and L-hercynine (HC) measurements

Plasma ET and HC were measured using liquid chromatography-tandem mass spectrometry (LC–MS/MS) as previously described [18]. Blood was collected from study participants into ethylenediaminetetraacetic acid (EDTA) tubes, centrifuged at 2000 *g* for 10 min at 4°C, then extracted for the upper plasma layers before storage at −80°C until use. ET, deuterated ET-d9, HC, and deuterated HC-d9 standards were obtained from ERGOLD (Paris, France). 10 μl of plasma was mixed with 105 μl methanol containing ET-d9 and HC-d9. Samples were vortexed and incubated at −20°C for 2h, followed by centrifugation and drying of supernatant under a stream of nitrogen gas. Dried samples were resuspended in 100 μl of ultrapure water and transferred to silanized vials for analysis. Liquid chromatography-tandem mass spectrometry (LC-MS/MS) was performed using an Agilent 1290 UPLC coupled to a 6460-QQQ mass spectrometer (Agilent Technologies, Santa Clara, CA, USA). Sample aliquots were kept at 10°C in the autosampler, then injected (2 μL) onto a Cogent Diamond-Hydride column (4 mm, 150 × 2.1 mm, 100 Å; MicroSolv Technology Corporation, Leland, NC, USA) at 30°C. The mobile phase consisted of 100% acetonitrile with 0.1% formic acid (solvent A) and 0.1% formic acid (solvent B). Chromatographic separation was achieved using a gradient elution at 0.4 ml/min from 25 % solvent B at 1 min to 40% solvent B over 3 min to elute ET (retention time of 3.6 min). MS was carried out under positive ion, electrospray ionization mode, using multiple reaction monitoring for quantification of specific target ions (see **Supplementary Table 2**). Capillary voltage was 3200 V, and gas temperature was maintained at 350°C. Nitrogen sheath gas pressure for nebulizing samples was maintained at 50 *psi*, with gas flow rate of 12.5 L/min. Ultra-high purity nitrogen was used as collision gas. Compounds were quantified with calibration standards (at least 8 concentrations encompassing the sample range), using the Agilent Mass Hunter software (Agilent Technologies), and subsequently dichotomized into Low or High ET, HC and HC:ET using their respective cohort-wide median values as cutoffs, which were 756.35nM, 11.54nM and 0.01417 respectively.

### 2.5. Statistical analysis

Statistical analysis was performed using R (version 4.4.0, R Core Team) and SPSS Statistics (v29, International Business Machines Corporation, Armonk, NY, USA)., Correlation analyses were performed using Spearman rank correlations, and group comparisons were performed using Mann-Whitney U tests due to the non-parametric distribution of several of the variables examined. For the primary outcome, we used linear regression to interrogate interaction effects of High vs. Low levels of plasma ET, HC or HC:ET on the association between plasma p-Tau and rate of cognitive decline (CDR-SB slope), where a significant interaction effect indicated that the strength of correlation between plasma p-tau and cognitive decline differs depending on the level of plasma ET, HC or HC:ET. For the secondary outcomes, Cox proportional hazards regression models were used to assess associations with incident cognitive and functional decline, where time to event was defined as the timepoint at which a participant’s CDR-GS had increased by ≥ 0.5 points compared to baseline. Linear mixed-effects (LME) models were used to examine associations with longitudinal cognitive performance, with global or domain-specific cognitive scores as outcome. The LME models contained random intercepts and random slopes, and time was modelled as a continuous variable. Time, groups (Group 1: Low A+ risk, Group 2: High A+ risk, Low HC:ET, Group 3: High A+ risk, High HC:ET) and the interaction between time and groups were included. All models were adjusted for age, sex, education, *APOE4* status, hypertension, hyperlipidaemia, diabetes, cardiovascular disease, and baseline CDR-SB. Analyses were performed separately for both plasma p-Tau181 and p-Tau217. Results were considered statistically significant at p<0.05.

## 3. Results

### 3.1. Participant characteristics

Demographic and clinical characteristics of the study cohort are listed in **Table 1**. A total of 259 dementia-free participants, including 77 (30%) NCI and 182 (70%) CIND, were included in the study. Participants were stratified into Low- (n=142, 55%), Intermediate- (n=67, 26%) or High- (n=50, 19%) risk of amyloid PET positivity (A+) using pre-determined plasma p-Tau181 reference ranges (**Supplementary Figure 2A**). Alternatively, in a subset of 255 participants with plasma p-Tau217 available, individuals were stratified into Low- (n=179, 70%), Intermediate- (n=10, 4%) or High- (n=66, 26%) risk of amyloid PET positivity (A+) using pre-determined plasma p-Tau217 reference ranges (**Supplementary Figure 2B**). Measures of longitudinal cognition include CDR-SB slope (change in CDR-SB per year, median [IQR], 0.045 [0.3]) and incident cognitive decline (increment in CDR-GS of ≥ 0.5, n=91, 35%), over a mean ± SD follow-up duration of 52 ± 15 months. To assess potential *APOE4* effects, ET, HC and HC:ET values were compared between *APOE4* carriers vs. non-carriers. **Supplementary Figure 3A** showed that ET, HC and HC:ET were not affected by APOE4 status in dementia-free participants (NCI + CIND) which constituted the current analyses for CR, while **Supplementary Figure 3B** showed reduced ET in the extant cohort including dementia group.

**Table 1.** Descriptive characteristics of the study cohort.

|  |  |
| --- | --- |
| **Study n** | 259 |
| Age, years, mean (SD) | 72 (8) |
| Female, n (%) | 129 (50) |
| Education, years, mean (SD) | 8 (5) |
| APOE ε4 carrier, n (%) | 66 (26) |
| Baseline CDR-SB, median (IQR) | 0 (1) |
| Baseline clinical diagnosis |  |
| NCI, n (%) | 77 (30) |
| CIND, n (%) | 182 (70) |
| **Vascular risk factors** |  |
| Hypertension, n (%) | 171 (66) |
| Hyperlipidemia, n (%) | 197 (76) |
| Diabetes, n (%) | 87 (34) |
| Cardiovascular disease, n (%) | 28 (11) |
| **Plasma biomarkers** |  |
| L-Hercynine : L-Ergothioneine ratio, median (IQR) | 0.012 (0.02) |
| L-Ergothioneine, nM, median (IQR) | 915 (955) |
| L-Hercynine, nM, median (IQR) | 13.0 (16) |
| **Plasma p-Tau181 derived reference ranges for identifying risk of amyloid PET positivity\*** |  |
| Low risk (Plasma p-tau181 ≤ 2.34pg/mL) | 142 (55) |
| Intermediate risk (Plasma p-tau181: 2.35pg/mL to 3.60pg/mL) | 67 (26) |
| High risk (Plasma p-tau181 ≥ 3.61pg/mL) | 50 (19) |
| **Plasma p-Tau217 derived reference ranges for identifying risk of amyloid PET positivity<sup>#</sup>** |  |
| Low risk (Plasma p-tau217 ≤ 11.39 NPQ) | 179 (70) |
| Intermediate risk (Plasma p-tau217: 11.40 to 11.53 NPQ) | 10 (4) |
| High risk (Plasma p-tau217 ≥ 11.54 NPQ) | 66 (26) |
| **Longitudinal cognitive performance** |  |
| CDR-SB slope, change/year, median (IQR) | 0.045 (0.3) |
| Increment in CDR-GS of ≥0.5, n (%) | 91 (35) |
| Follow-up time, months, mean (SD) | 52 (15) |

\* Risk stratified using pre-determined plasma p-Tau181 reference ranges (**Supplementary Figure 2A**). Plasma p-Tau181 measured at either baseline visit (n=245) or 12-month follow up visit (n=14).
<sup>#</sup> Risk stratified using pre-determined plasma p-Tau217 reference ranges (**Supplementary Figure 2B**). Plasma p-Tau217 available in 255 participants and were measured at either baseline visit (n=206), 12-month (n=47) or 24-month (n=2) follow up visit. **Abbreviations:** CDR-SB, Clinical Dementia Rating Sum of Boxes; CDR-GS, Clinical Dementia Rating Global Score; CIND, Cognitive Impairment No Dementia; IQR, Interquartile Range; NCI, No Cognitive Impairment; PET, positron emission tomography; SD, Standard Deviation

### 3.2. Associations of plasma p-Tau with brain amyloid and cognitive decline

Plasma p-Tau181, as an established marker of amyloid pathology, expectedly correlated with PET amyloid marker Pittsburgh compound B’s standardized uptake value ratio (PiB-PET SUVr, rho=0.653, p<0.001; **Figure 1A**). Higher levels of p-Tau181 also correlated with faster cognitive decline (as measured by the CDR-SB slope, **Figure 1C**). As another biomarker, p-Tau217 has been reported to have superior utility as a marker of amyloid pathology, especially the early pathological transition from amyloid accumulation to tau hyperphosphorylation and dysregulation [27, 29], we also analyzed plasma p-Tau217 measured on the NULISA platform, and confirmed that it significantly correlated with PiB-PET SUVr (**Figure 1B**), and CDR-SB slope (**Figure 1D**). Taken together, our data support the use of p-Tau181 and p-Tau217 as markers of brain amyloid pathology.

**Figure 1.**
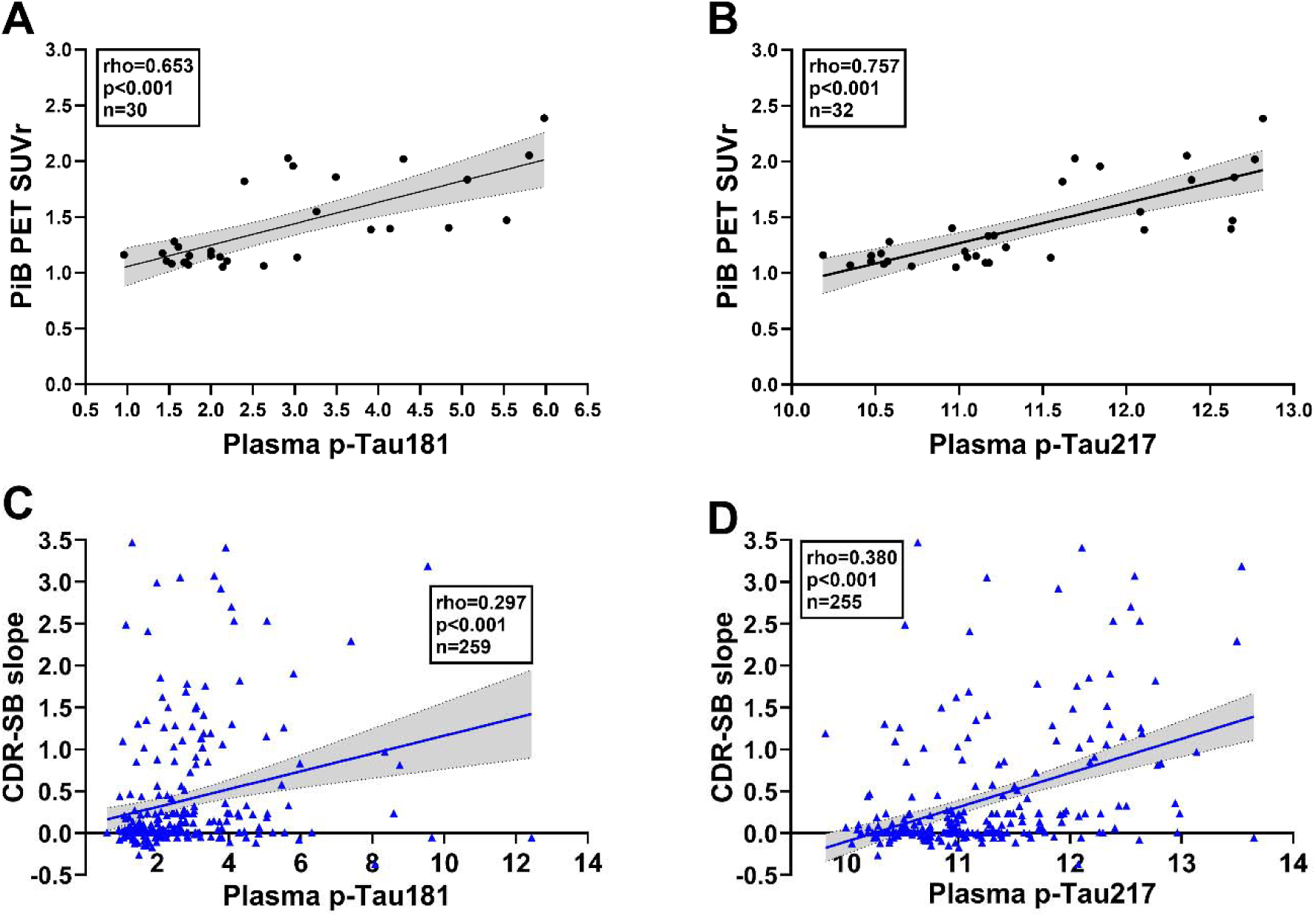
Associations of plasma p-Tau with brain amyloid and cognitive decline. Graphs showing Spearman’s correlations of available PiB-PET SUVr (as a measurement of cortical amyloid burden) with plasma (**A**) p-Tau181 or (**B**) p-Tau217; as well as correlations of CDR-SB slope (change in CDR-SB per year, as a measurement of cognitive decline) with plasma (**C**) p-Tau181 or (**D**) p-Tau217. Solid line indicates linear regressed best-fit curve while dashed lines indicate 95% confidence intervals. Unit for plasma p-Tau181 = pg/mL while unit for plasma p-Tau217 = NPQ. ***Abbreviations:*** *CDR-SB, Clinical Dementia Rating Sum of Boxes; NPQ, NULISA protein quantification units; PiB-PET, Pittsburgh Compound B - positron emission tomography; p-Tau181, phosphorylated Tau181; p-Tau217, phosphorylated Tau217; SUVr, Standardized uptake value ratio*.

### 3.3. Association between plasma p-Tau levels and cognitive decline is moderated by L-hercynine to L-ergothioneine ratio

By themselves, higher L-Hercynine (HC), L-ergothioneine (ET) as well as their ratio (HC:ET) were weakly correlated with lower cognitive decline (rho from −0.134 to −0.293, see **Supplementary Figure 4**). To investigate whether ET metabolism may have stronger effects on the associations between p-Tau levels and cognitive decline, we performed moderation analyses based on High vs Low HC:ET. **Figure 2A** shows that interaction between HC:ET with log-transformed plasma p-Tau181 levels remained significant (P_interaction_=0.042) even after adjusting for covariates. Specifically, HC:ET moderated the association between plasma p-Tau181 levels and cognitive decline, such that participants with High HC:ET showed slower cognitive decline with increasing plasma p-Tau181 (β=0.0976; 95% CI= −0.444 to 0.639); while those with Low HC:ET showed the expected strong positive correlation between plasma p-Tau181 and cognitive decline (β=0.849;95% CI= 0.318 to 1.38). In contrast, no significant interactions were observed for ET (P_interaction_=0.937) and HC (P_interaction_=0.182, data not shown). Repeating the analyses with non-log-transformed plasma p-Tau181 showed similar effects (**Supplementary Figure 5**). Consistent results were obtained for p-Tau217, with a strong moderation effect by HC:ET (P_interaction_=0.004, see **Figure 2B**).

**Figure 2.**
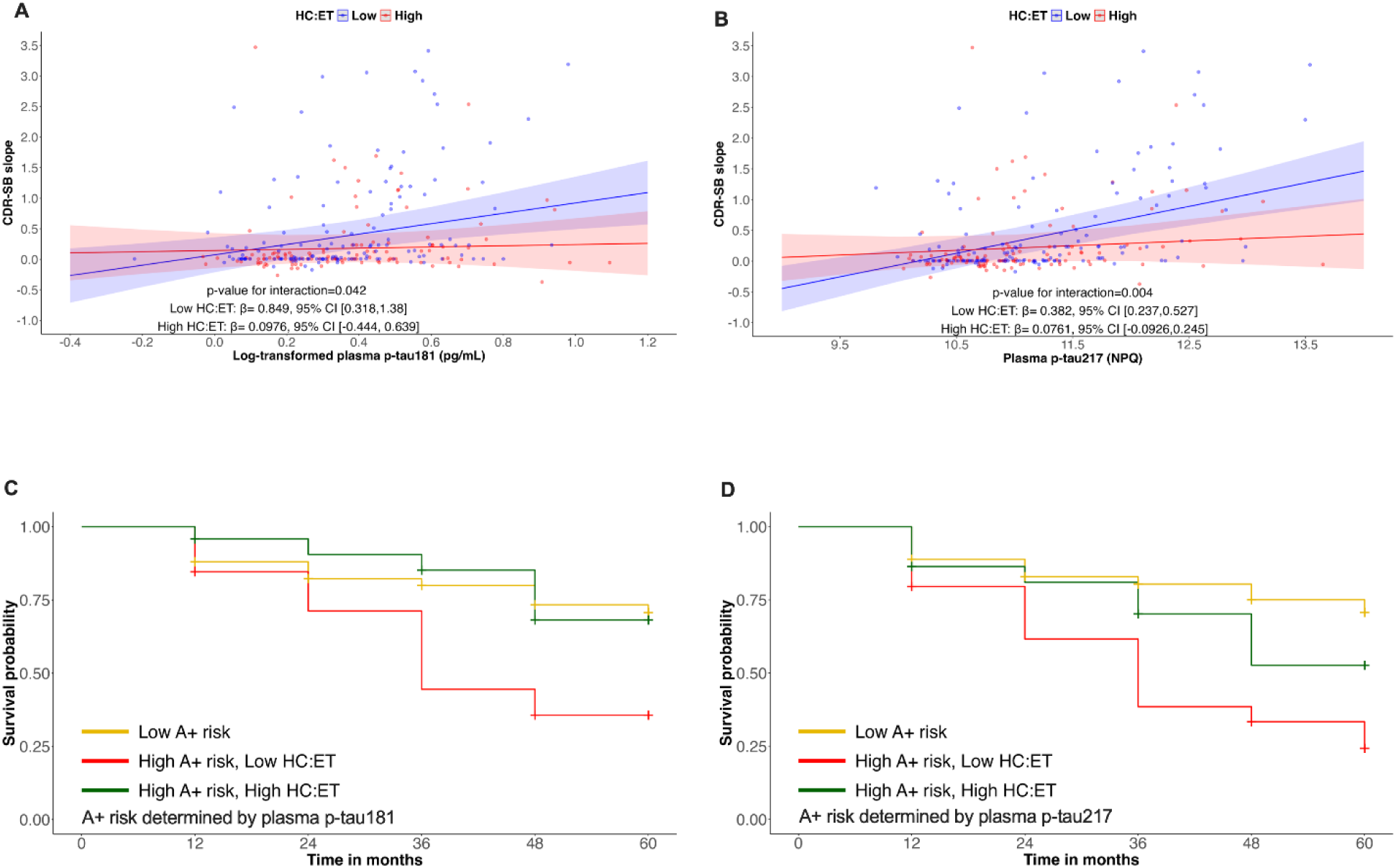
Associations between plasma p-Tau and cognitive decline in participants stratified by plasma L-hercynine to L-ergothioneine ratio. The associations between plasma (**A**) p-Tau181 (log-transformed) or (**B**) p-Tau217 and CDR-SB slope (change in CDR-SB per year, as a measure of cognitive decline), is weaker in participants with High plasma HC:ET (pink lines) compared to Low HC:ET (purple lines), with High vs. Low HC:ET dichotomized using median values. Shaded areas around each regression line represent the 95% confidence intervals for the regression lines. Regression analyses were adjusted for age, sex, education, *APOE4* status, hypertension, hyperlipidaemia, diabetes, cardiovascular disease and baseline CDR-SB. (**C**) and (**D**) are Kaplan-Meier curves for participants stratified by Low- (yellow lines) vs. High-Risk of amyloid PET positivity (A+) based on plasma p-Tau181 (see **Supplementary Figure 2A**) and p-Tau217 (see **Supplementary Figure 2B**) respectively, with the High-Risk group further stratified by Low (red lines) vs. High (green lines) HC:ET. Incident cognitive decline is defined as an increment of CDR-GS of ≥ 0.5 from baseline. Unit for plasma p-Tau181 = pg/mL while unit for plasma p-Tau217 = NPQ. ***Abbreviations:*** *CDR-GS, Clinical Dementia Rating Global Scores; CDR-SB, Clinical Dementia Rating Sum of Boxes; ET, plasma L-ergothioneine concentration; HC, plasma L-hercynine concentration; NPQ, NULISA protein quantification units; p-Tau181, phosphorylated Tau181; p-Tau217, phosphorylated Tau217*.

### 3.4. Risk of incident cognitive decline in participants at high risk of amyloid PET positivity is influenced by L-hercynine to L-ergothioneine ratio

Next, we used plasma p-Tau181 cut-offs to identify participants at Low- (plasma p-Tau181 ≤ 2.34pg/mL) or High-Risk (plasma p-Tau181 ≥ 3.61pg/mL) for amyloid PET positivity (A+). A CDR-GS increase of ≥ 0.5 used to define risk of incident cognitive decline has previously been shown to be clinically relevant in detecting cognitive decline and progression to AD [23, 30]. Cox regression analyses (**Table 2A**) of participants with High-Risk for A+ showed that those with Low HC:ET had an approximately twofold increased risk of incident cognitive decline over the follow-up period (Hazard ratio [HR] =1.96; 95% Confidence Interval [CI] =1.01 to 3.79; p=0.047) compared to those with Low-Risk for A+. In contrast, no increased risk was observed in High A+ participants with High HC:ET (HR= 0.87; 95% CI=0.33 to 2.26; p=0.770). Similar results were observed when applying plasma p-Tau217 cut-offs to identify individuals at Low-(plasma p-Tau217 ≤ 11.39 NPQ) or High-Risk (plasma p-Tau217 ≥ 11.54 NPQ) for A+. Compared to participants with Low-Risk for A+, those with High-Risk for A+, Low HC:ET were associated with higher risk of incident cognitive decline (HR =2.33; 95% CI =1.38 to 3.93; p=0.002; see **Table 2B**). Again, this was not observed in High A+, High HC:ET participants (HR= 1.47; 95% CI=0.69 to 3.10; p=0.318). **Figure 2C** and **Figure 2D** depict the corresponding Kaplan-Meier survival curves for p-Tau181 and p-Tau217, respectively. Comparisons of plasma p-Tau and HC:ET levels across the three groups (Low A+ risk, High A+ risk with Low HC:ET, and High A+ risk with High HC:ET) are presented in **Supplementary Figure 6**, which showed that both plasma p-Tau181 and p-Tau217 levels were significantly higher in High A+ risk vs. compared to Low A+ risk, but did not differ between High A+ with Low HC:ET vs. High A+ with High HC:ET. Conversely, mean HC:ET values were higher in the High vs. Low HC:ET subgroups within the High A+ risk groups. Taken together, these results support the clinical relevance of analyses based on dichotomized variables.

**Table 2.** Cox regression analyses of risk for cognitive decline in participants stratified by plasma p-Tau and L-hercynine to L-ergothioneine ratio.

**(A) Risk of amyloid PET positivity (A+) determined by plasma p-Tau181**
| Cognitive decline defined as an increment of CDR-GS of $\geq 0.5$ (Outcome) | | | | | |
| --- | --- | --- | --- | --- | --- |
| Risk of amyloid PET positivity<br>(determined by plasma p-tau181)<br>and plasma HC:ET status (n=192) | Stable,<br>n (%) | Declined,<br>n (%) | Hazard<br>ratio | 95% CI | p-<br>value |
| Low-Risk for A+ (n=142) | 103 (73) | 39 (27) |  | Reference |  |
| High-Risk for A+, Low HC:ET (n=26) | 11 (42) | 15 (58) | **1.956** | **1.010, 3.788** | **0.047** |
| High-Risk for A+, High HC:ET (n=24) | 18 (75) | 6 (25) | 0.867 | 0.333, 2.257 | 0.770 |

**(B) Risk of amyloid PET positivity (A+) determined by plasma p-Tau217**
| Cognitive decline defined as an increment of CDR-GS of $\geq 0.5$ (Outcome) | | | | | |
| --- | --- | --- | --- | --- | --- |
| Risk of amyloid PET positivity<br>(determined by plasma p-tau217)<br>and plasma HC:ET status (n=245) | Stable,<br>n (%) | Declined,<br>n (%) | Hazard<br>ratio | 95% CI | p-<br>value |
| Low-Risk for A+ (n=179) | 131 (73) | 48 (27) |  | Reference |  |
| High-Risk for A+, Low HC:ET (n=44) | 14 (32) | 30 (68) | **2.328** | **1.380, 3.928** | **0.002** |
| High-Risk for A+, High HC:ET (n=22) | 13 (59) | 9 (41) | 1.465 | 0.692, 3.100 | 0.318 |

### 3.5. Trajectories of global and domain-specific cognitive performance in participants at high risk of amyloid PET positivity are influenced by L-hercynine to L-ergothioneine ratio

Within the group identified as High-Risk for A+ (using plasma p-Tau181 cut-offs), those with Low HC:ET showed significantly faster decline in global cognition (**Supplementary Figure 7A**) and in all cognitive domains except attention (**Supplementary Table 3A**), compared with the Low-Risk for A+ group. In contrast, the High-Risk for A+, High HC:ET group exhibited faster decline only in executive function and memory, with changes in the other cognitive domains not significantly different from those of the Low A+ risk group. Corresponding analyses using plasma p-Tau217 cut-offs showed that the High A+ risk / Low HC:ET group declined faster in global cognition as well as across all cognitive domains (**Supplementary Figure 7B** and **Supplementary Table 3B**). Similar results were obtained for the High A+ risk / High HC:ET group except for the attention and language cognitive domains, which were not significantly different from those of the Low A+ risk group. Taken together, these results suggest that among individuals at high risk for amyloid pathology, having high ET metabolism was associated with less extensive and slower cognitive decline compared to those with low ET metabolism.

## 4. Discussion

The concept of cognitive resilience as the ability of individuals to maintain brain cognition and function amid aging and disease is gaining increasing momentum in AD research, due to the potential for further insights into neuroprotective mechanisms which can counter AD processes, as well as advancing processes and molecules as potential therapeutic targets. In this context, it is important to recognize that cognitive resilience is a general term which includes cognitive reserve (CR), brain reserve (BR) and brain maintenance (BM), each of which refers to specific aspects of resilience mechanisms, either acquired or innate [2, 3]. In this longitudinal cohort study, we provide the first clinical evidence that increased L-hercynine to L-ergothioneine ratio (HC:ET) attenuated the cognitive decline typically observed in older people with high risk of brain amyloid positivity, and was also associated with lower risk of incidental cognitive decline for up to five years. These findings indicate that plasma HC:ET may be a novel biomarker of cognitive resilience in AD. Oxidative stress has been recognized as both promoter [31] and consequence [32] of amyloid pathology, and is associated with mitochondrial dysfunction, glucose hypometabolism and neuronal death [33]. L-Ergothioneine (ET), a diet-derived thione, has originally been characterized as a potent antioxidant [13] whose effects are putatively mediated via its oxidative desulfurization to L-hercynine (HC) by reactive oxygen species (ROS), in effect rendering ET an important ROS scavenger [14, 15]. Therefore, HC:ET may be interpreted as an index of ET metabolism, and our results suggest that efficient scavenging and countering ROS-associated neurotoxic effects may be one mechanism by which high ET metabolism confers resilience to amyloid pathology-associated cognitive decline.

### 4.1. Unanswered questions and future work

Although the current findings suggest that ET-associated antioxidant effects may contribute to cognitive resilience, a major limitation of this interpretation is the failure of supplementation with common antioxidants like vitamin E in AD interventional trials [34, 35]. Postulated underlying reasons for the lack of efficacy include inadequate target engagement (as brain levels of antioxidants remained low), complex pathophysiology involving multiple processes besides oxidative stress, intervention occurring too late as well as insufficient dosing and duration [35–37]. For some of these drawbacks, ET may offer advantages due to its unique pharmacokinetic properties. For example, the neurophysiological importance of ET is supported by findings that its cognate transporter, organic cation transporter 1 (OCTN1, encoded by *SLC22A4*), is highly expressed in brain parenchyma, accounting for high levels of distribution to neural cells [38, 39] leading to improved target engagement. Furthermore, ET is differentiated from other antioxidants pharmacodynamically by manifesting a wide range of beneficial effects besides antioxidation, leading them to be called a conditionally essential micronutrient [40], nutraceutical [41], or even a longevity vitamin [42]. Examples of putative neuroprotectant ET effects which may counter AD pathophysiological processes include anti-neuroinflammation, promotion of neuronal differentiation, upregulation of neurotrophic factors, increased dendritic spine formation and inhibition of β-amyloid oligomer formation [14, 39, 42, 43]. However, it is unclear which of the aforementioned mechanism(s) mediate cognitive resilience, and which underlying components (CR, BR or BM) may be involved. Indeed, some purported mechanisms may impinge on more than one component. For example, the upregulation of neurotrophic factors may affect CR by increasing efficiency of neural networks, but may also involve BR by stimulating synapse structural integrity, as well as BM by maintaining synaptic plasticity processes. Therefore, further work is needed to comprehensively characterize ET metabolism and associated effects which underlie the different components of cognitive resilience.

A second major unanswered question relate to our findings of significant moderation effects for HC:ET. This suggests that, in contrast to dementia where ET deficits were evident [18], the baseline efficiency of ET metabolism is a more important factor in the case of cognitive resilience. However, the biological factors regulating HC:ET, including functional polymorphic variants of OCTN1, or the potential involvement of gut microbiome known to alter absorption or uptake [34], are at present unknown, and should be further studied to better assess the potential utility of ET-based therapeutic strategies.

Finally, the finding of reduced ET in *APOE4* carriers in participant groups which include those with dementia (see **Supplementary Figure 3B**) is in agreement with our previous study showing reduced ET in AD dementia [18]. However, it is currently unclear whether the association between ET and *APOE4* carrier status is indirect (i.e., ET is disease stage-dependently related to AD dementia, which is in turn associated with higher prevalence of *APOE4*), or there may be a direct pathophysiological link between *APOE4* and ET absorption or transport. Follow-up mechanistic studies would be worthwhile.

### 4.2. Other Limitations

Whilst plasma phosphorylated tau species, p-Tau181 and p-Tau217 have been well-validated as peripheral markers of cortical amyloid burden [25, 44] (also see **Figure 1**), more established measurements using amyloid PET or cerebrospinal fluid tests were not performed for the entire cohort. Furthermore, while amyloid plaques are the hallmark neuropathological feature of AD which facilitate the emergence and spread of tau [45], clinically it is phosphorylated-tau-associated neurofibrillary tangle (NFT) burden [45] and potentially certain types of Aβ oligomers [46, 47] which more closely associate with neurodegeneration and cognitive decline. Future studies should therefore examine whether HC:ET moderates the associations of NFT or Aβ oligomer measures (both clinical and postmortem) with longitudinally assessed cognitive decline. On the other hand, we have previously demonstrated the potential clinical utility of plasma p-Tau by showing increased levels predict for faster cognitive decline [27, 48]. Thus, our current study add to the clinical relevance of plasma p-Tau217 and p-Tau181 by showing that among individuals at high risk of plasma p-Tau-defined amyloid positivity, those with High HC:ET had a mitigated risk of cognitive decline (**Figure 2** and **Table 2**), indicating plasma index of ET metabolism as a novel biomarker of cognitive resilience to amyloid pathology. This finding warrants validation in larger, more diverse cohorts to assess generalizability.

Finally, whilst we did not measure dietary intake of ET in the current clinical study, we have previously found, in a separate non-dementia cohort, that compared with eating mushrooms (a major source of ET) less than once per week, consuming more than two portions per week was associated with lower risk of mild cognitive impairment even after adjusting for demographic, vascular, and lifestyle covariates [49]. This suggests that intervention with dietary ET might have beneficial effects on CR [50]. However, it is worth noting that our current study found associations with HC:ET, and not ET *per se*, so other parameters related to ET metabolism may be important besides ET concentrations. Perhaps ET supplementation may be more beneficial for individuals whose ET levels are in deficit, while for others with normal ET levels, factors effecting ET metabolism might be more critical. Future investigations with careful measurements of ET intake or large randomized controlled trials with ET supplementation may help address these questions.

## 5. Conclusion

A plasma index of L-ergothioneine metabolism may be a novel biomarker of cognitive resilience to amyloid pathology, indicating the need for further investigations of underlying mechanisms which may involve both well-known antioxidant effects as well as emergent neuroprotective properties. L-Ergothioneine metabolism-associated effects, as well as factors regulating the efficiency of L-ergothioneine metabolism, should be further assessed as potential therapeutic targets for cognitive decline and dementia.

## Declaration of the use of generative AI and AI-assisted technologies in scientific writing and in figures, images and artwork

The authors declare that no generative AI or AI-assisted technologies were used in the writing of this manuscript or in the preparation of figures, images, or artwork.

## Ethics statement

Approval for the study was obtained from the Singapore National Healthcare Group Domain-Specific Review Board (2018/00996, 2015/00406, and 2015/00441). Written informed consent was obtained from study participants or their authorized next of kin.

## Data availability

Anonymized datasets generated during and/or analyzed in the current study are available from the corresponding author upon reasonable request.

## CRediT authorship contribution statement

**Joyce R. Chong:** Writing – review & editing, Writing – original draft, Conceptualization, Methodology, Investigation, Formal analysis, Visualization, Data curation. **Irwin Cheah:** Writing – review & editing, Methodology, Investigation, Formal analysis. **Richard Tang:** Writing – review & editing, Methodology, Investigation. **Barry Halliwell:** Writing – review & editing, Methodology. **Christopher P. Chen:** Writing – review & editing, Data curation, Supervision, Funding acquisition. **Mitchell K. P. Lai:** Writing – review & editing, Conceptualization, Visualization, Data curation, Supervision, Project administration, Funding acquisition.

## Acknowledgments

We are grateful to the patients and their families for their participation in this study. We acknowledge the coordinator and rater teams from the Memory, Ageing and Cognition Centre for assistance with participant recruitment and assessment. Joyce Chong is a recipient of an A*STAR International Fellowship from the Agency for Science, Technology and Research (A*STAR) Singapore, which played no role in the writing of this manuscript or the decision to publish.

## Supplementary materials

- Supplementary Figure 1. Flow chart of study cohort
- Supplementary Figure 2. Three-range reference for amyloid PET positivity using plasma p-Tau181 and p-Tau217 concentrations
- Supplementary Figure 3. Comparison of plasma HC, ET, and HC:ET levels between *APOE4* non-carriers and carriers
- Supplementary Figure 4. Correlations of plasma biomarkers with cognitive decline
- Supplementary Figure 5. Associations between plasma p-Tau181 and cognitive decline in participants stratified by plasma L-hercynine to L-ergothioneine ratio
- Supplementary Figure 6. Comparisons of plasma p-Tau and HC:ET levels across Low A+ risk, High A+ risk with Low HC:ET, and High A+ risk with High HC:ET groups
- Supplementary Figure 7. Trajectories of cognitive decline in participants stratified by plasma p-Tau and plasma L-hercynine to L-ergothioneine ratio (global cognition)
- Supplementary Table 1. Neuropsychological battery and component tests
- Supplementary Table 2. Mass spectrometry multiple reaction monitoring parameters
- Supplementary Table 3. Trajectories of cognitive decline in participants stratified by plasma p-tau and plasma L-hercynine to L-ergothioneine ratio (global and domain-specific cognitive tests)

