## Supplemental for "Metabolism of the antioxidant micronutrient ergothioneine as a plasma biomarker of cognitive resilience in older people with Alzheimer’s disease amyloid pathology"

### Supplementary Materials

#### Supplementary Figure 1. Flow chart of study cohort

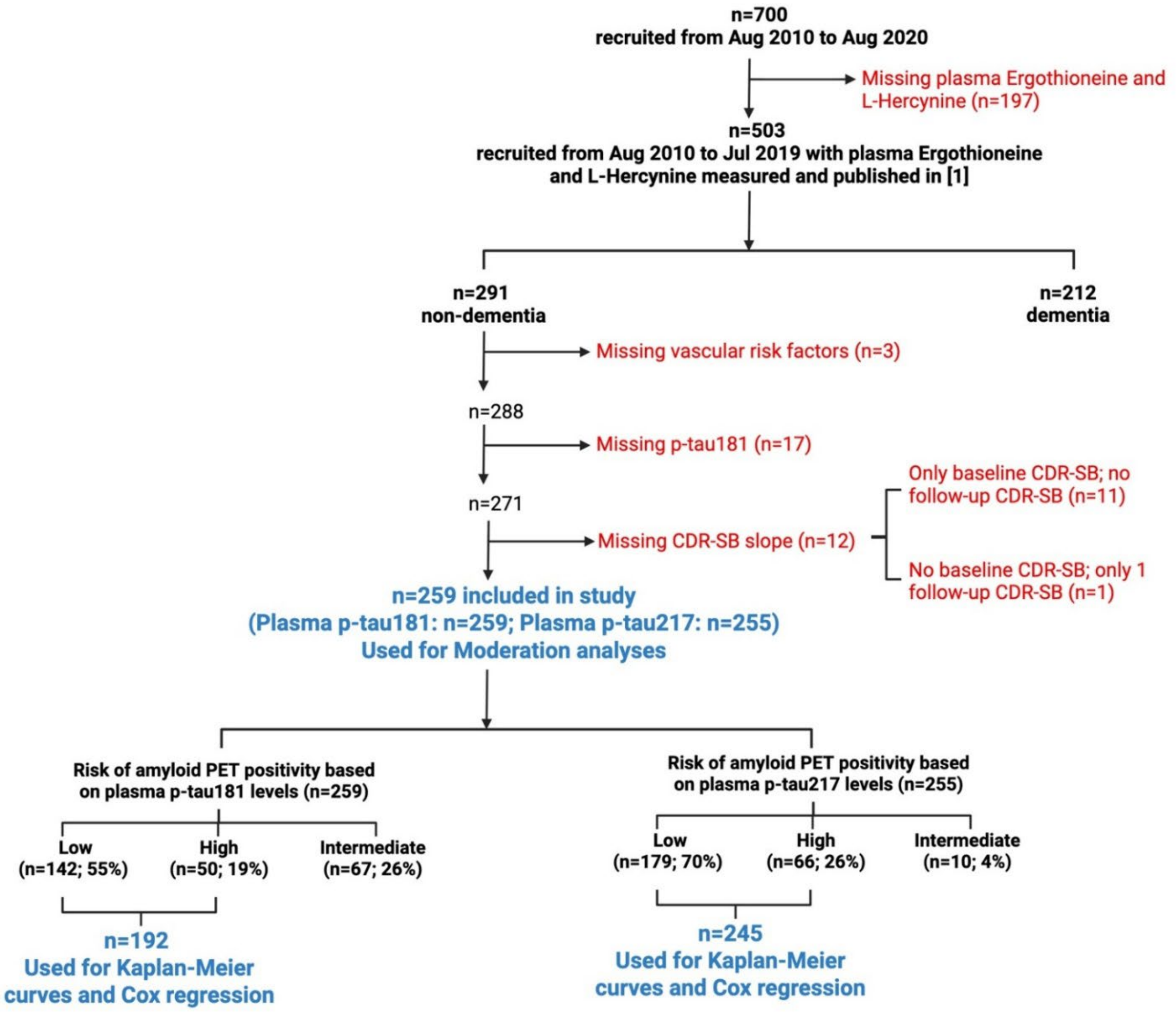


Of the 700 participants recruited between August 2010 and August 2020, 503 (recruited from August 2010 to July 2019) had both plasma L-ergothioneine (ET) and L-hercynine (HC) levels measured, as previously published [3]. Among these 503 participants, 291 were diagnosed as dementia-free (NCI or CIND). Of the dementia-free participants, 32 were excluded due to missing data: 3 lacked data on vascular risk factors, 17 lacked plasma p-Tau181, and 12 did not have at least two CDR-SB assessments required to determine the CDR-SB slope (change in CDR-SB per year). As a result, 259 participants with complete data required for moderation analyses were included in this study. Of the 259 participants, plasma ET, HC and p-Tau181 were measured at either baseline visit (n=245) or 12-month follow up visit (n=14). All participants had CDR-SB recorded at baseline as well as at least one annual follow-up visit. Using pre-determined plasma p-Tau181 reference ranges (see **Supplementary Figure 2A**), participants were stratified into low- (n=142), high- (n=50) and intermediate- (n=67) risk of amyloid PET positivity. Plasma p-Tau217 was available in a subset of 255 participants. Using pre-determined plasma p-Tau217 reference ranges (see **Supplementary Figure 2B**), participants were stratified into low- (n=179), high- (n=66) and intermediate- (n=10) risk of amyloid PET positivity. Participants with low- or high-risk of amyloid PET positivity were included in the Kaplan-Meier curves and Cox regression analyses. ***Abbreviations:*** *CDR-SB, Clinical Dementia Rating Sum of Boxes; PET, Positron Emission Tomography*

**Supplementary Figure 2. Three-range reference for amyloid PET positivity using plasma p-Tau181 and p-Tau217 concentrations**

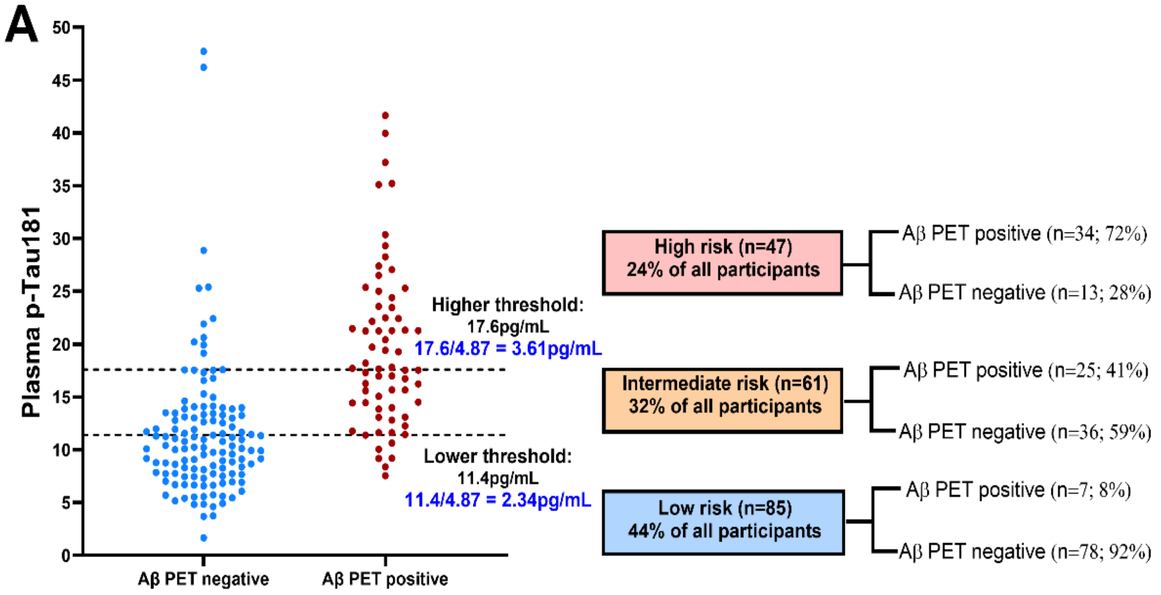


| Plasma p-Tau181 cut-offs (pg/mL) in amyloid PET cohort | Plasma p-Tau181 cut-offs (pg/mL) in current study cohort (factor of 4.87 applied) | Risk group |
|---|---|---|
| ≥17.6 | ≥3.61 | High |
| 11.5 to 17.5 | 2.35 to 3.60 | Intermediate |
| ≤11.4 | ≤2.34 | Low |

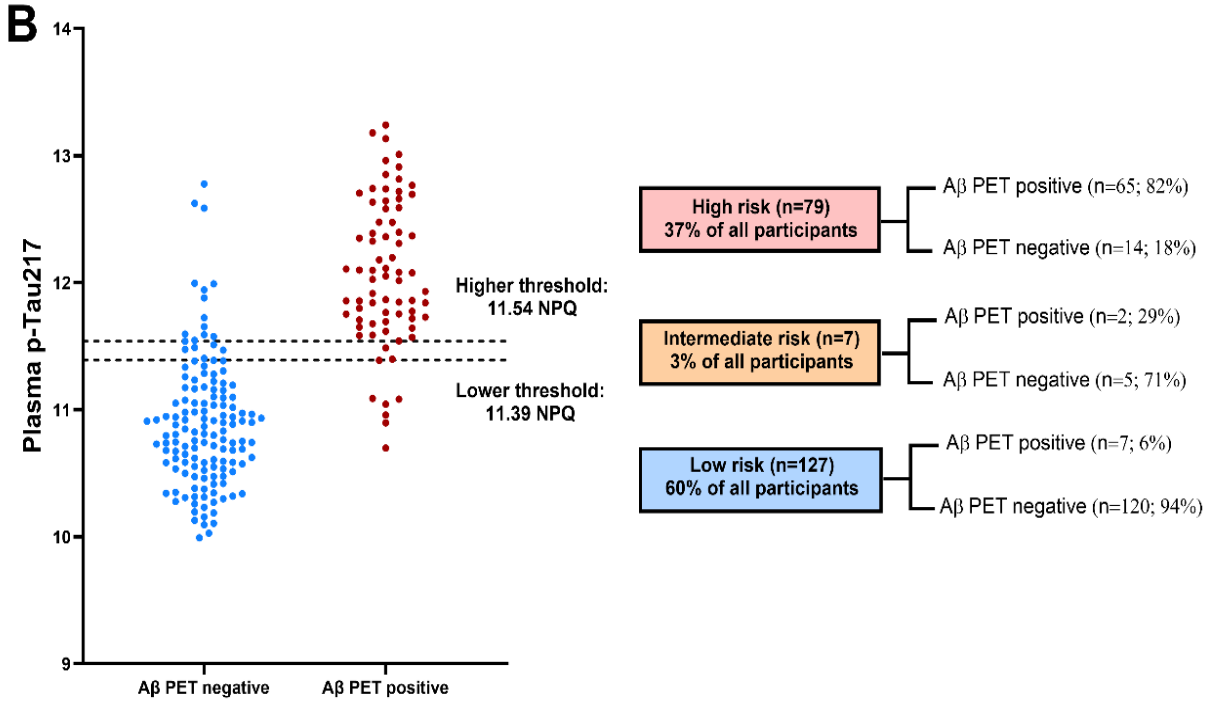

**Supplementary Figure 2.** Amyloid PET procedure to measure brain amyloid burden for the cohort has been previously published [1]. Briefly, imaging was conducted using [$^{11}$C]Pittsburgh Compound B (PiB) or [$^{18}$F]flutafuranol amyloid tracer radioligands. All images were reconstructed using ordinary Poisson ordered-subsets expectation maximization with all corrections applied. Global standardized uptake values ratio (SUVr) were derived from the [$^{11}$C]PiB scans and individual parcellated MRI for reference and target region definition using an inhouse developed automated pipeline [2]. Each [$^{11}$C]-PiB or [$^{18}$F]-flutafuranol image was independently visually interpreted by three experienced raters blinded to the clinical diagnosis of each participant, and was used to classify individuals as Aβ positive (Aβ+) or Aβ negative (Aβ-) following established criteria. To pre-determine the reference ranges for Aβ-PET positivity, a local cohort with both plasma p-Tau181 and amyloid PET data available was used. The average time between blood collection and time of PET is 0.2 months (range 0 - 9.2 months). A three-range strategy was applied, comprising a lower reference point to rule out AD (90% sensitivity; low risk for PET Aβ+) and a higher reference point to rule in AD (90% specificity; high risk for PET Aβ+). (**A**) The graph shows the distribution of plasma p-Tau181 concentrations. The blue dots corresponded to individuals who are Aβ-PET negative and red dots to individuals who are Aβ-PET positive. The lower dashed line demonstrates where the 90% sensitivity low-risk threshold falls on the distribution, with the upper line corresponding to the 90% specificity high-risk threshold. On the right of the graph, the flowchart demonstrated the overall accuracy of the workflow, when intermediate-risk individuals are referred to confirmatory Aβ-PET testing, for predicting Aβ-PET positivity based on the 90% Sensitivity/Specificity strategy. As different versions of Simoa p-Tau181 assays were used in the amyloid PET cohort (Simoa p-Tau181 assay developed in University of Gothenburg) and current study (Simoa® pTau-181 Advantage V2 Kit [item 103714]), normalization was performed. Briefly, matched plasma samples (n=29) from the amyloid PET cohort were analyzed in the current study. High concordance (Spearman's rho=0.768, $p<0.001$) was achieved between the samples despite the lower absolute values of the current study. Based on this data, a correction factor of 4.87 was applied to the thresholds derived in the amyloid PET cohort to adjust to the current study cohort. The adjusted thresholds are shown in **blue** font. Units for plasma p-Tau181 = pg/mL. A separate set of analyses was also done using plasma p-Tau217, where the average time between blood collection and time of PET is 35 months (range 0 - 97 months) using a similar three-range strategy. (**B**) The graph shows the distribution of plasma p-Tau217 measurements using the NULISA™ platform (Alamar Biosciences, Fremont, CA, USA). The blue dots corresponded to individuals who are Aβ-PET negative and red dots to individuals who are Aβ-PET positive. The lower dashed line demonstrates where the 90% sensitivity low-risk threshold falls on the distribution, with the upper line corresponding to the 90% specificity high-risk threshold. On the right of the graph, the flowchart demonstrated the overall accuracy of the workflow for predicting Aβ-PET positivity based on the 90% Sensitivity/Specificity strategy. Units for plasma p-Tau217 = NPQ. ***Abbreviations:*** *Aβ-PET, amyloid positron emission tomography; NPQ, NULISA Protein Quantitation units*

**Supplementary Figure 3. Comparison of plasma HC, ET, and HC:ET levels between *APOE4* non-carriers and carriers**

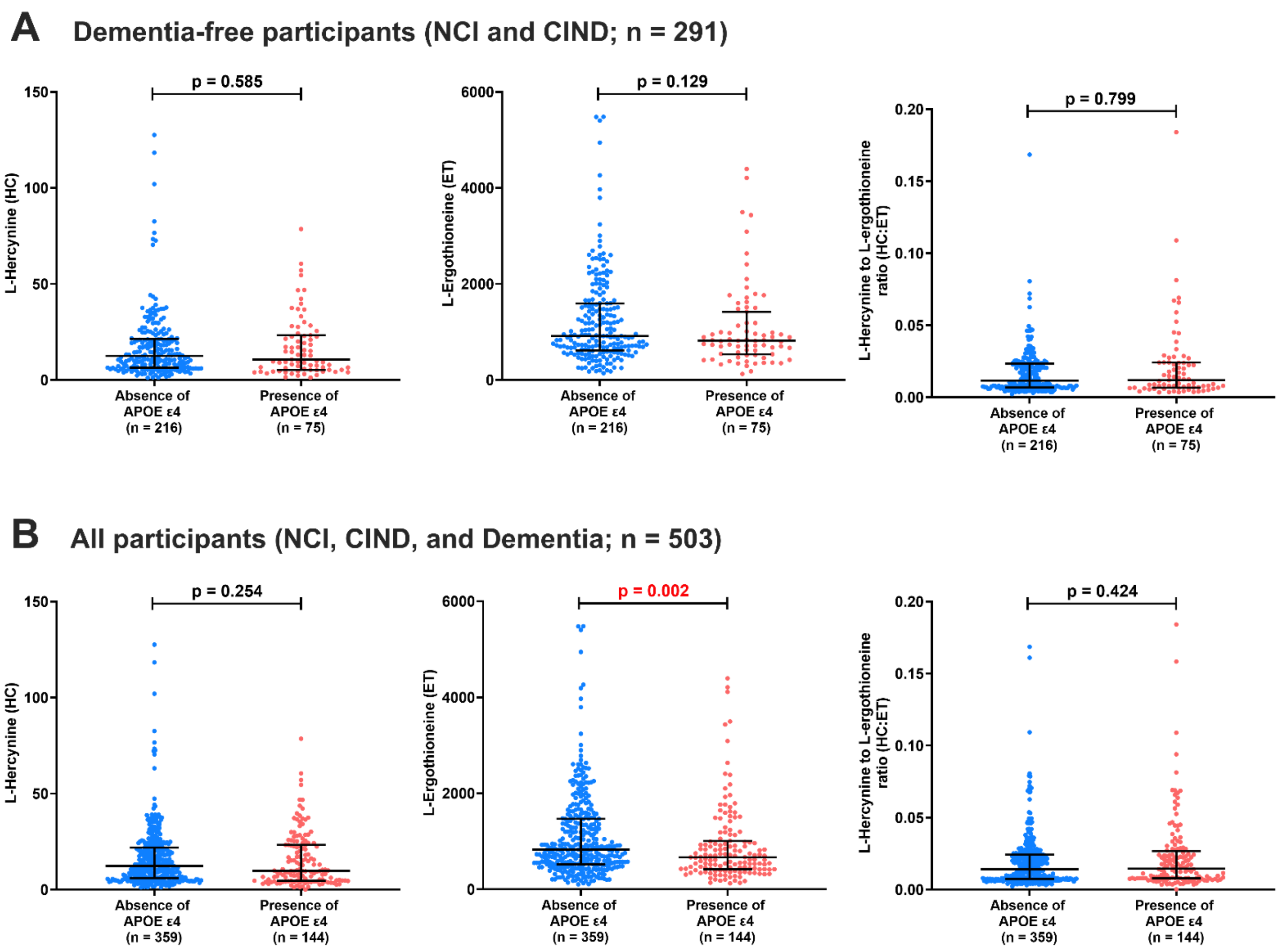


Comparison of plasma HC, ET, and HC:ET levels between *APOE4* non-carriers ("Absence of APOE ε4") and carriers ("Presence of APOE ε4") among (**A**) dementia-free participants (NCI and CIND), and (**B**) all participants, including those with NCI, CIND, and dementia. The graphs show the median and interquartile range. p-values derived from Mann-Whitney U test. **Red** indicates significant p-values ($p < 0.05$). For better visualization of the graphs, one participant in the *APOE4* non-carrier group was not displayed in the ET graphs (plasma ET =8133.21nM). Units for HC and ET = nM. ***Abbreviations:*** *CIND, Cognitive Impairment No Dementia; ET, L-ergothioneine; HC, L-Hercynine; NCI, No Cognitive Impairment*

**Supplementary Figure 4. Correlations of plasma biomarkers with cognitive decline**

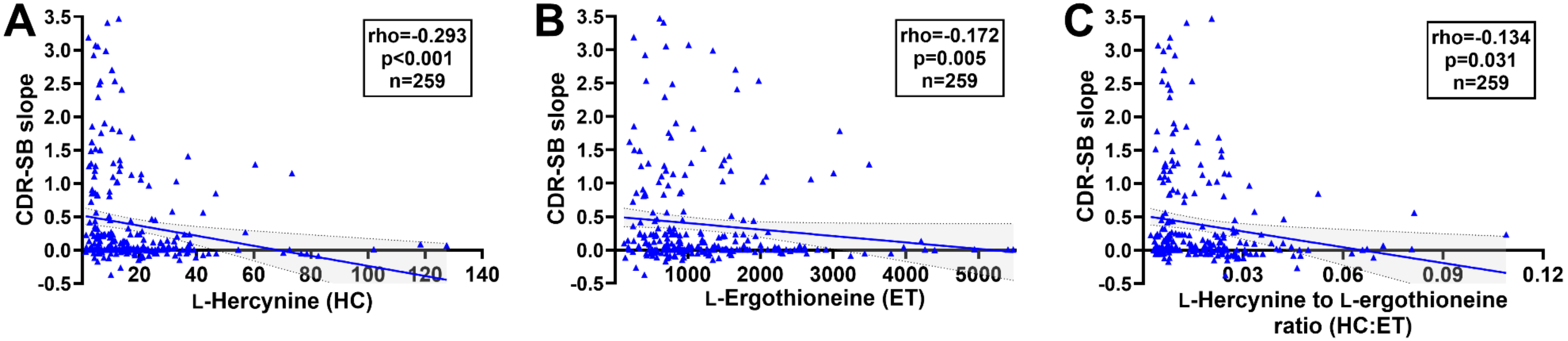


Spearman's correlations between plasma (**A**) HC; (**B**) ET and (**C**) HC:ET and CDR-SB slope (change in CDR-SB / year) as a measurement of cognitive decline. Solid line indicates linear regressed best-fit curve while dashed lines indicate 95% confidence intervals. For better visualization of the graphs, one participant was excluded from the graph in (B) (plasma ET = 8133.21nM, CDR-SB slope = 0.089), while two participants were excluded from the graph in (C) (HC:ET = 0.169, CDR-SB slope = 0.14; plasma HC:ET = 0.184, CDR-SB slope = -0.054). Units for HC and ET = nM. ***Abbreviations:*** *CDR-SB, Clinical Dementia Rating Sum of Boxes; ET, L-ergothioneine; HC, L-Hercynine.*

**Supplementary Figure 5. Associations between plasma p-Tau181 and cognitive decline in participants stratified by plasma L-hercynine to L-ergothioneine ratio**

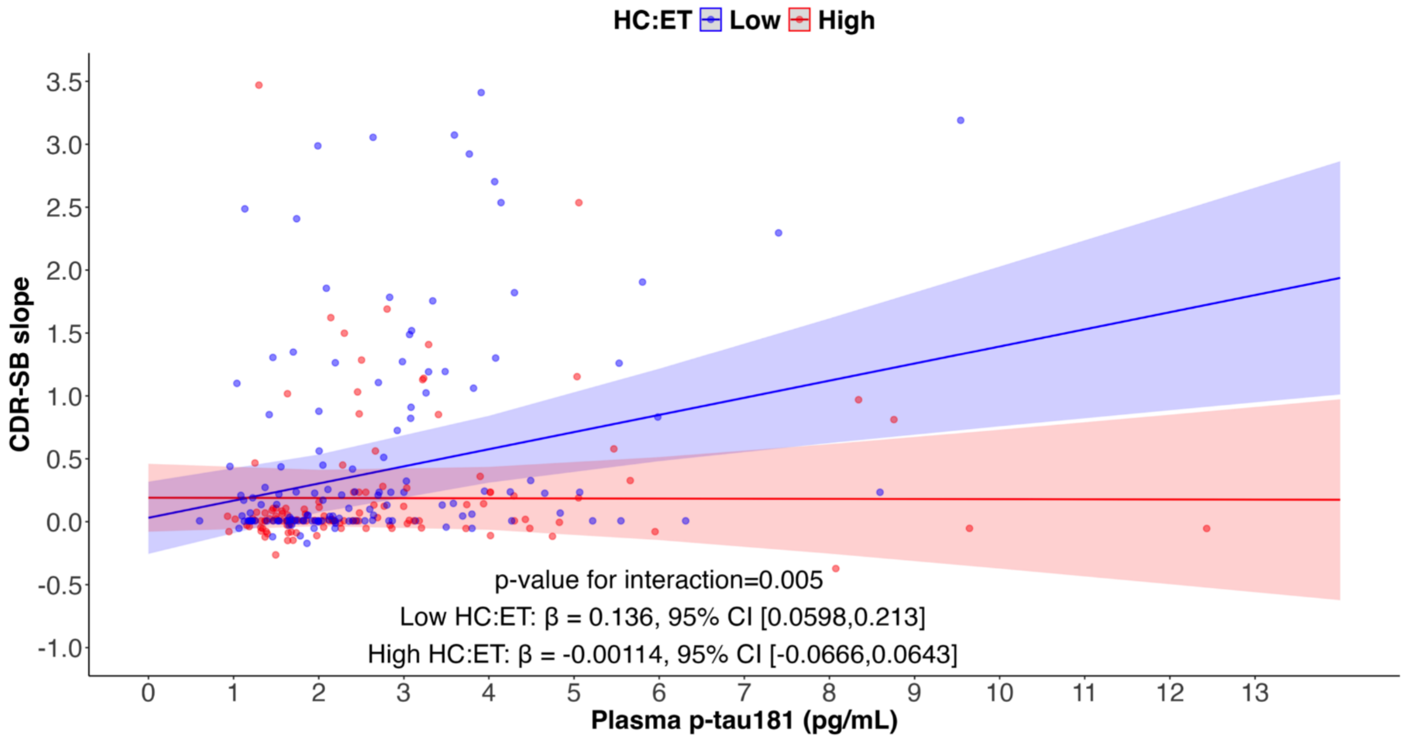


The association between plasma p-Tau181 (non-log-transformed) and CDR-SB slope (change in CDR-SB per year, as a measure of cognitive decline), is weaker in participants with High plasma HC:ET (pink lines) compared to Low HC:ET (purple lines). High vs. Low HC:ET was dichotomized using the median value. Shaded areas around each regression line represent the 95% confidence intervals for the regression lines. Regression analyses were adjusted for age, sex, education, APOE Ɛ4 status, hypertension, hyperlipidemia, diabetes, cardiovascular disease and baseline CDR-SB. Units for plasma p-Tau181 = pg/mL. ***Abbreviations:*** *CDR-SB, Clinical Dementia Rating Sum of Boxes; ET, plasma L-ergothioneine concentration; HC, plasma L-hercynine concentration; NPQ, NULISA Protein Quantitation units*

**Supplementary Figure 6. Comparisons of plasma p-Tau and HC:ET levels across Low A+ risk, High A+ risk with Low HC:ET, and High A+ risk with High HC:ET groups**

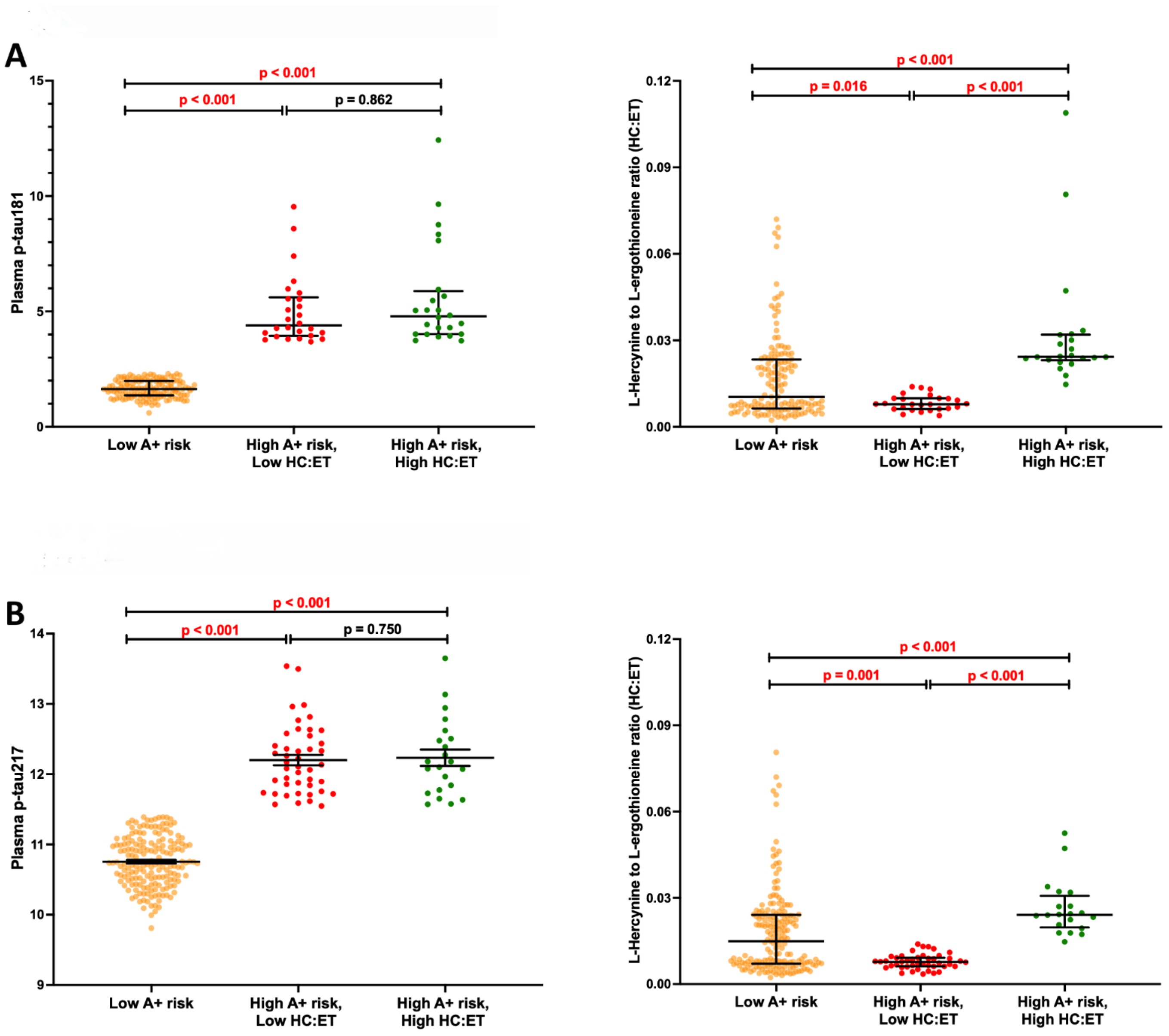


Comparisons of respective plasma p-Tau levels and HC:ET amongst brain amyloid positivity risk (A+) groups based on cut-off values of (**A**) plasma p-Tau181 and (**B**) plasma p-Tau217. The graphs show the median and interquartile range for plasma p-Tau181 and HC:ET, and the mean and standard error of the mean for plasma p-Tau217. p-values derived from Kruskal–Wallis tests followed by *post hoc* Dunn's tests for plasma p-Tau181 and HC:ET, and one-way ANOVA followed by *post hoc* least significant difference (LSD) tests for plasma p-Tau217. **Red** indicates significant p-values ($p < 0.05$). For better visualization of the graphs, two participants in the High A+ risk, High HC:ET group were not displayed in the HC:ET graphs (HC:ET = 0.169 and 0.184, respectively). Units for plasma p-Tau181 = pg/mL; plasma p-Tau217 = NPQ; ET and HC = nM. ***Abbreviations:*** *ANOVA, Analysis of variance; ET, Plasma L-ergothioneine concentration; HC, Plasma L-hercynine concentration; HC:ET, Ratio of plasma L-hercynine to L-ergothioneine; NPQ, NULISA Protein Quantitation units.*

**Supplementary Figure 7. Trajectories of cognitive decline in participants stratified by plasma p-Tau and plasma L-hercynine to L-ergothioneine ratio (global cognition)**

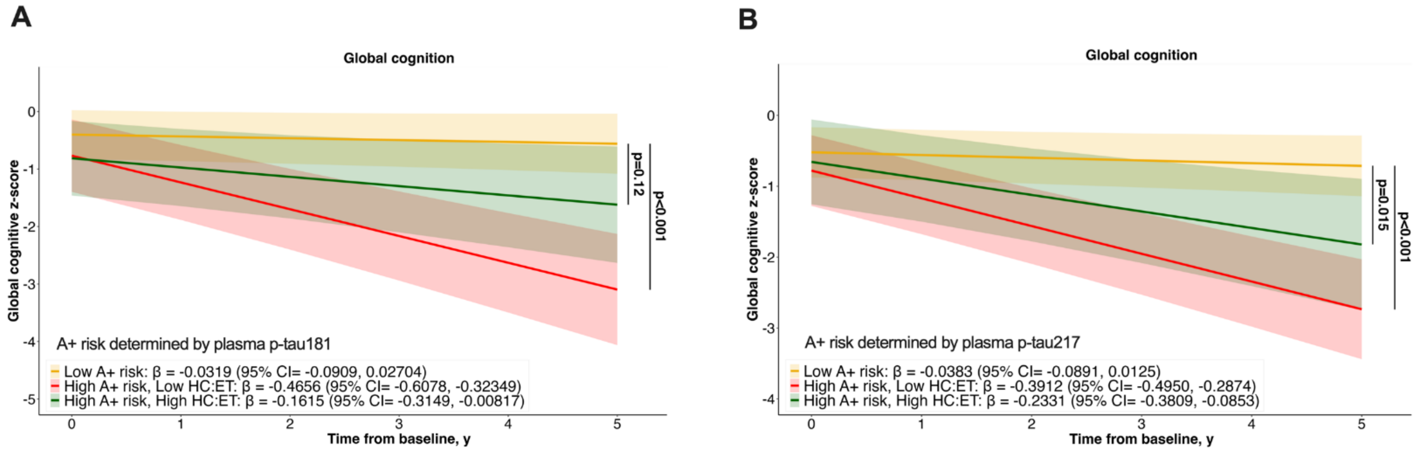


Trajectories of global cognition in participants stratified by Low- vs. High-risk of amyloid PET positivity (A+), with the High-risk group further stratified by plasma levels of HC:ET (Low vs. High). Risk of amyloid PET positivity was derived based on plasma p-Tau181 (**A**; refer to **Supplementary Figure 2A** for the plasma p-Tau181 cut-offs) or plasma p-Tau217 (**B**; refer to **Supplementary Figure 2B** for the plasma p-Tau217 cut-offs). Estimated means of global cognitive *z*-scores with 95% CI are presented. β-coefficients (95% CI) were derived from linear mixed-effect models adjusted for age, sex, education, *APOE* ε4 status, hypertension, hyperlipidaemia, diabetes, cardiovascular disease, and baseline CDR-SB. p-values indicate the statistical significance of differences in rates of change in global cognitive z-score when compared to the Low A+ Risk group. ***Abbreviations:*** *CDR-SB, Clinical Dementia Rating Sum of Boxes; ET, Plasma L-ergothioneine concentration; HC, Plasma L-hercynine concentration; HC:ET, Ratio of plasma L-hercynine to L-ergothioneine*

**Supplementary Table 1. Neuropsychological battery and component tests**

| Cognitive Domain | Component Test(s) |
|---|---|
| **Executive Function:** | Frontal Assessment Battery [3] |
| **Attention:** | Digit Span, Visual Memory Span [4] and Auditory Detection [5] |
| **Language:** | Modified Boston Naming Test [6] and Verbal Fluency [7] |
| **Visuomotor Speed:** | Symbol Digit Modality Test [8, 9], Maze Task [9] and Digit Cancellation [10] |
| **Visuoconstruction:** | Weschler Memory Scale – Revised (WMS-R) Visual Reproduction Copy task [4], Clock Drawing [11] and Weschler Adult Intelligence Scale – Revised (WAIS-R) subtest of Block Design [12] |
| **Visual Memory:** | Picture Recall & Recognition Tasks, and WMS-R Visual Reproduction Recall & Recognition Task [4] |
| **Verbal Memory:** | Word List Recall & Recognition Tasks [13] and Story Recall Task |

Table shows the component neuropsychological tests of seven cognitive domains used in a comprehensive battery to categorize clinical subgroups. All individual test raw scores were transformed to standardized *z*-scores using the means and standard deviation (SD) of the non-cognitively impaired (NCI) group. The score for each domain was created by averaging the *z*-scores of individual component tests and standardized using the composite mean and SD of the NCI group. To obtain the global cognition score for each patient, the domain *z*-scores were averaged and standardized using the mean and SD of the NCI group. At follow-up, global and domain-based cognitive *z*-scores were obtained using the means and SDs of the NCI group at baseline.

**Supplementary Table 2. Mass spectrometry multiple reaction monitoring parameters**

| Target | Precursor ion | Product ion | Fragmentor voltage (V) | Collision energy (eV) |
|---|---|---|---|---|
| **ET** | 230 | 186 | 103 | 9 |
| **ET-$d_9$** | 239 | 195 | 98 | 9 |
| **HC** | 198 | 95 | 94 | 21 |
| **HC-$d_9$** | 207 | 95 | 103 | 9 |

ET = L-ergothioneine; ET-$d_9$ = Deuterated L-ergothioneine; HC = L-hercynine; HC-$d_9$ = Deuterated L-hercynine

**Supplementary Table 3. Trajectories of cognitive decline in participants stratified by plasma p-Tau and plasma L-hercynine to L-ergothioneine ratio (global and domain-specific cognitive tests)**

| | Risk of A+ based on plasma p-Tau181 | | |
|---|---|---|---|
| **A** | **Low A+ risk β (95% CI)** | **High A+ risk, Low HC:ET β (95% CI)** | **High A+ risk, High HC:ET β (95% CI)** |
| **Global cognition** | -0.0319 (-0.0909, 0.02704) | -0.4656 (-0.6078, -0.32349)* | -0.1615 (-0.3149, -0.00817) |
| **Executive function** | 0.024 (-0.0214, 0.0694) | -0.286 (-0.3962, -0.1762)* | -0.109 (-0.2286, 0.0104)* |
| **Attention** | -0.0428 (-0.0805, -0.00505) | -0.1282 (-0.2195, -0.03686) | -0.0859 (-0.1853, 0.01355) |
| **Language** | -0.0394 (-0.171, 0.0916) | -0.8878 (-1.204, -0.5714)* | -0.1508 (-0.493, 0.1912) |
| **Visuospatial function** | -0.0712 (-0.118, -0.0239) | -0.2358 (-0.350, -0.1213)* | -0.1232 (-0.248, 0.0016) |
| **Visuomotor speed** | -0.0328 (-0.055, -0.0106) | -0.1634 (-0.217, -0.1096)* | -0.0870 (-0.146, -0.0282) |
| **Memory** | 0.0578 (0.0204, 0.0953) | -0.1467 (-0.2373, -0.0560)* | -0.1194 (-0.2181, -0.0207)* |

| | Risk of A+ based on plasma p-Tau217 | | |
|---|---|---|---|
| **B** | **Low A+ Risk β (95% CI)** | **High A+ Risk, Low HC:ET β (95% CI)** | **High A+ Risk, High HC:ET β (95% CI)** |
| **Global cognition** | -0.0383 (-0.0891, 0.0125) | -0.3912 (-0.4950, -0.2874)* | -0.2331 (-0.3809, -0.0853)* |
| **Executive function** | 0.0105 (-0.0313, 0.0523) | -0.2497 (-0.3356, -0.1639)* | -0.1374 (-0.2592, -0.0157)* |
| **Attention** | -0.0388 (-0.0702, -0.00739) | -0.1430 (-0.2077, -0.07832)* | -0.0550 (-0.1466, 0.03647) |
| **Language** | -0.046 (-0.157, 0.0645) | -0.667 (-0.894, -0.4411)* | -0.231 (-0.552, 0.0910) |
| **Visuospatial function** | -0.0673 (-0.111, -0.0236) | -0.2205 (-0.310, -0.1306)* | -0.2141 (-0.341, -0.0868)* |
| **Visuomotor speed** | -0.0387 (-0.0582, -0.0193) | -0.1378 (-0.1778, -0.0978)* | -0.1006 (-0.1573, -0.0440)* |
| **Memory** | 0.0552 (0.0232, 0.0872) | -0.1202 (-0.1860, -0.0545)* | -0.1775 (-0.2707, -0.0842)* |

Trajectories of cognitive decline in participants stratified by Low- vs. High-Risk of amyloid PET positivity (A+), with the High-Risk group further stratified by plasma levels of HC:ET (Low vs. High). Risk of amyloid PET positivity was derived based on plasma p-Tau181 (**A**; refer to **Supplementary Figure 2A** for the plasma p-Tau181 cut-offs) or plasma p-Tau217 (**B**; refer to **Supplementary Figure 2B** for the plasma p-Tau217 cut-offs). Estimated means of global/domain-specific cognitive z-scores with 95% CI are presented. β-coefficients (95% CI) were derived from linear mixed-effect models adjusted for age, sex, education, *APOE* ℇ4 status, hypertension, hyperlipidaemia, diabetes, cardiovascular disease, and baseline CDR-SB. *Indicates significantly different rates of change in cognitive scores when compared to the Low A+ risk group at $p < 0.05$. ***Abbreviations:*** *ET, Plasma L-ergothioneine concentration; HC, Plasma L-hercynine concentration*
